# Antibody responses to AZD1222 vaccination in West Africa

**DOI:** 10.1101/2022.05.04.22274668

**Authors:** Adam Abdullahi, David Oladele, Steven A. Kemp, James Ayorinde, Abideen Salako, Fehintola Ige, Douglas Fink, Chika Onwuamah, Qosim Osuolale, Rufai Abubakar, Azuka Okuruawe, Gideon Liboro, Oluwatosin Odubela, Gregory Ohihoin, Oliver Ezechi, Olagoke Usman, Sunfay Mogaji, Adedamola Dada, Soraya Ebrahimi, Lourdes Ceron Gutierrez, Sani H. Aliyu, Rainer Doffinger, Rosemary Audu, Richard Adegbola, Petra Mlcochova, Babatunde Lawal Solako, Ravindra K. Gupta

## Abstract

**Background:** There are no real world data on vaccine elicited neutralising antibody responses for the world’s most widely used vaccine, AZD1222, in African populations following scale up. Here, we measured **i)** baseline SARS-CoV-2 seroprevalence and levels of protective neutralizing antibodies prior to vaccination rollout using both flow cytometric based analysis of binding antibodies to nucleocapsid (N), coupled with virus neutralisation approaches and **ii)** neutralizing antibody responses to VOC prior to vaccination (January 2021) and after two-doses of AZD1222 vaccine administered with a 12 week interval in Lagos, Nigeria - a period when the Delta variant was circulating.

**Methods:** Health workers at multiple sites in Lagos were recruited to the study. For binding antibody measurement, IgG antibodies against SARS-COV-2 Wuhan-1 receptor-binding domain (RBD), trimeric spike protein (S), nucleocapsid protein (N) and Omicron S1 were measured using the Luminex-based SARS-CoV-2-IgG assay by flow cytometry. For plasma neutralising antibody measurement, SARS-CoV-2 lentiviral pseudovirus (PV) were prepared by transfecting 293T cells with Wuhan-614G wild type (WT), B.1.617.2 (Delta) and BA.1 (Omicron) plasmids in conjunction with HIV-1 expression vectors and luciferase encoding genome flanked by LTRs. We performed serial plasma dilutions from each time point and mixed plasma with PV before infecting HeLa-ACE2 cell lines, reading out luminescence and calculating ID50 (reciprocal dilution of sera required to inhibit 50% of PV infection).

**Results:** Our underlying study population receiving at least one dose of vaccine comprised 140 participants with a median age of 40 (interquartile range: 33, 48). 62/140 (44%) participants were anti-N IgG positive prior to administration of first vaccine dose. 49 had plasma samples available at baseline prior to vaccination and at two follow-up timepoints post two dose vaccination for neutralization assays. Half of the participants, 25/49 (51%) were IgG anti-N positive at baseline. Of the 24 individuals anti-N Ab negative at baseline, 12/24 had ID_50_ above the cut-off of 20. In these individuals, binding antibodies to S were also detectable, and neutralisation correlated with IgG anti-S, suggesting waning of N antibody after infection. Overall, neutralizing Ab titres to WT 1 month after second dose were 2579 and at 3 months post second-dose were 1695. As expected, lower levels of neutralization were observed against the Delta GMT 549 and Omicron variants 269 at 1 month. Positive anti-N IgG Ab status at baseline was associated with significantly higher titres of neutralizing antibodies following vaccination across all tested VOC. Those with anti-N Abs present at baseline did not experience waning of responses between months 1 and 3 post second dose. When data were analysed for negative anti-N IgG status at any timepoint, there was a significant decline in neutralization and binding antibodies between 1 month and 3 months post second-dose. The GMT in these individuals for Delta and Omicron was approximately 100, nearly a log lower in comparison to WT. We tested anti-N IgG in subjects who were anti-N IgG negative at baseline (n=78) and became positive between 1- and 3-months post second dose and found 7/49 (14%) with *de-novo* infection, with one additional participant demonstrating both reinfection and breakthrough infection to yield a total breakthrough rate of 8/49 (16%). Neutralising and binding Ab titres 1 month post vaccine, prior to breakthrough, were not associated with breakthrough infection. Neutralizing titres were higher at the last time point in individuals who had experienced vaccine breakthrough infection (with no evidence of infection prior to vaccine), indicating a boosting effect of infection in addition to vaccine. However, neutralisation and binding S antibodies against Omicron were low in those with either prior exposure or infection following two dose AZD1222.

**Conclusions:** AZD1222 is immunogenic in this real world west African cohort with significant background seroprevalence and incidence of breakthrough infection over a short time period. Prior infection and breakthrough infection induced higher anti-SARS-CoV-2 Ab responses at 3 months post vaccine against all widely circulating VOC. However, responses to Omicron BA.1 were low at three months regardless of hybrid immunity from prior exposure or breakthrough infection. Booster doses after AZD1222 should be considered in the African setting, even after natural infection, as future variants may be more pathogenic as well as immune evasive in the context of waning immunity.

## Introduction

AZD1222 is a chimpanzee adenovirus-vectored vaccine (ChAdOx1) based on the SARS-CoV-2 spike protein. Adenoviral vectored vaccines generally generate lower neutralising antibody responses compared to mRNA vaccines^1-4^. T cell responses across both platforms appear to be robust and well preserved, as is protection from severe disease and death^5-7^.

Despite highly effective vaccines, SARS-CoV-2 transmission continues. Variants of concern (VOC), likely arising within chronically infected patients^8^ demonstrating both immune escape and increased infectivity ^2,3,9-13^ have compromised protective effects of two dose vaccines such as BNT162b2 and AZD1222 in the context of suboptimal vaccine coverage/waning of immune responses^3^. Although a third dose with mRNA vaccination is able to rescue neutralisation against B.1.529 Omicron in the short term^3,12^, waning has been documented in vulnerable individuals. Fourth doses increase immune responses and are being implemented in some countries for higher risk populations^14^.

Across the African region, vaccine rollout has been heterogenous with 16% of total eligible population completing vaccination and 1.3% receiving a booster dose^1^. Serum neutralization in vitro correlates with protection against SARS-COV-2 infection in clinical studies^15^. However, vaccine rollout in African countries is impaired by paucity of neutralisation data and vaccine efficacy data for VOC; in particular, there are no data on vaccine elicited neutralising antibody responses for AZD1222-the world’s most widely used vaccine-in African populations following scale up. These data are particularly important given the continual emergence of new immune evasive variants.

Here, we measured **i)** baseline SARS-CoV-2 seroprevalence and levels of protective antibodies prior to vaccination rollout using both flow cytometric based analysis of binding antibodies to nucleocapsid (N), coupled with virus neutralisation approaches and **ii)** neutralizing antibody responses to VOC prior to vaccination (January 2021) and after two-doses of AZD1222 vaccine administered between June and July 2021 in Lagos, Nigeria, during a period when the Delta variant was also circulating.

## Methods

### Study population and sampling

Health care workers (HCWs) and Health workers (HWs) at the Nigerian Institute of Medical Research (NIMR) and Federal Medical Center, Ebute Metta, volunteering to be vaccinated with two doses of AZD1222 twelve weeks apart were recruited to the study following signed informed consent. HCWs were defined as patient facing staff such as nursing and midwifery professionals, pharmacist, social workers, and laboratory scientists involved in nasopharyngeal sample collection. HWs were defined as non-patient facing staff such as computing professionals, administrative associate professionals, secretaries, clerks, drivers and laboratory scientist involved in sample processing. The study design comprised a prospective longitudinal cohort study of adult patients who were eligible to receive their first dose vaccination between 13 March 2021 and 31 March 2021 and prospectively recruited into the NIMR vaccine effectiveness study. Participants provided plasma sample at baseline (prior to first-dose, T0), before second dose (T1), 1 month after second dose (T2) and 3 months after second dose (T3) [Figure 1].

**Figure 1:**
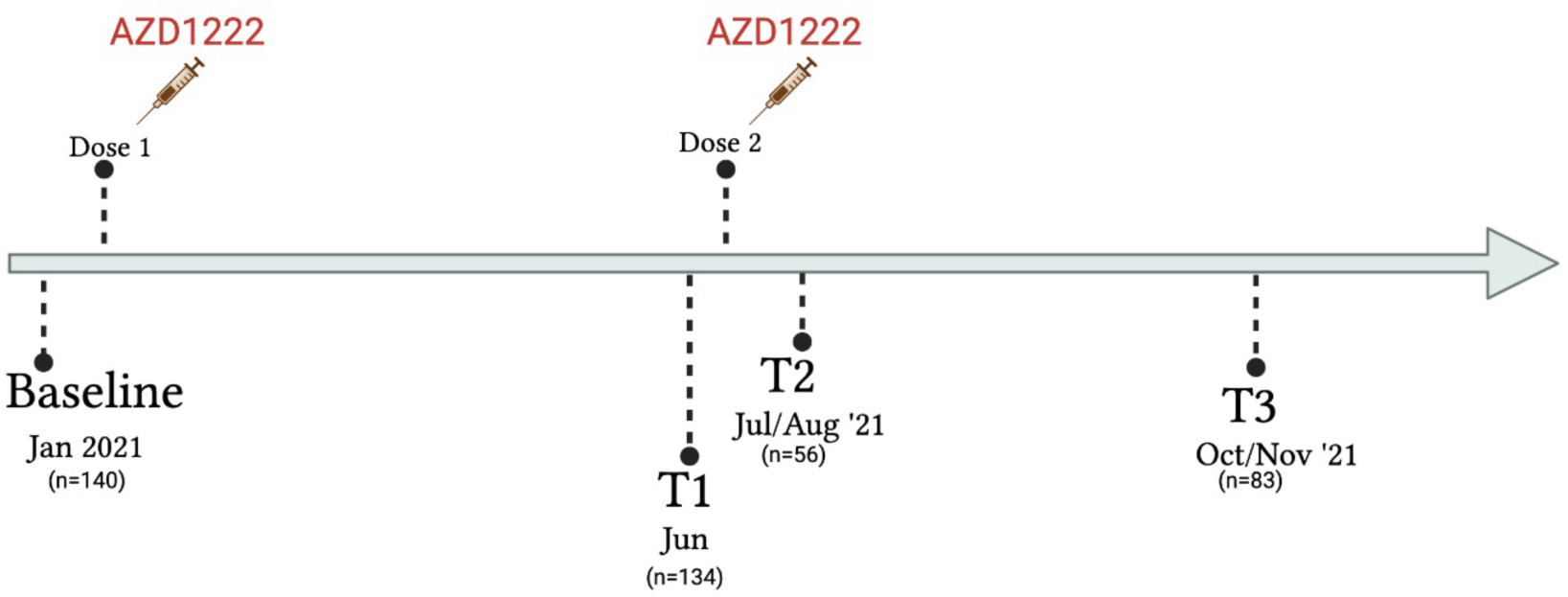
Study design and flow of patient disposition for recipients of AZD1222 two dose vaccination.

### Laboratory methods and sample testing

Binding IgG antibodies (Abs) against SARS-COV-2 receptor-binding domain (RBD), trimeric spike protein (S) and nucleocapsid protein (N) were measured using the Luminex-based SARS-CoV-2-IgG assay (Luminex) by flow cytometry as previously detailed^8,16^. We defined previous SARS-CoV-2 infection as positive anti-N IgG above cut-off of 6104 mean fluorescence intensity (MFI), based on analysis of ‘true’ positive (convalescent) and negative pre pandemic samples. Vaccine breakthrough infection was defined as the absence of IgG anti-N at 1-month post-second dose and its presence at 3-months post second-dose.

For plasma neutralising antibody measurement, SARS-CoV-2 lentiviral pseudovirus (PV) were prepared by transfecting 293T cells with Wuhan-614G wild type (WT), B.1.617.2 (Delta) and BA.1 (Omicron) plasmids in conjunction with p8.91 HIV-1 gag-pol expression vector ^17^. We and others previously showed high correlation between PV and live virus neutralisation^18,19^. Sample testing was performed at baseline, T2 and T3 timepoints. Pseudovirus neutralization was performed on Hela-ACE2 cells using SARS-CoV-2 spike PV expressing luciferase. Briefly, plasma samples were heat inactivated at 54°C for 1 hour, serially diluted in duplicates and incubated with PVs at 37°C for 1 hour prior to addition of Hela-ACE2 cells^4^. Plasma dilution/virus mix were incubated for 48 hours at 5% CO_2_ environment at 37°C, and luminescence was measured using Bright-Glo Luciferase assay system (Promega). All neutralization assays were repeated in two independent experiments containing 2 technical replicates for each condition. Neutralization was calculated relative to virus-only controls as a mean neutralization with s.e.m. Half maximum inhibitory dose (ID_50_) was calculated in GraphPad Prism version 9.0 and ID_50_>20 was considered positive.

### Statistical analysis

Geometric Mean Titre (GMT) with 95% confidence interval (CI) of neutralization antibody were calculated across timepoints. Characteristics of participants were expressed as proportions and percentages for categorical variables and median inter quartile range (IQR) for continuous variables. Mann-Whitney or Wilcoxon test was used to compare neutralization antibody titres across timepoints and compare participants based on IgG anti-N strata. Differences between neutralization antibody titres in IgG anti-N participants with ID_50_>20 were compared by Kruskal Wallis non-parametric test. Statistical analysis was performed using GraphPad Prism version 9.0.

### Ethics

This study was approved by the Institutional Review Board of NIMR (IRB-21-040). All participants gave written informed consent.

## Results

### Evidence of prior SARS-COV-2 infection by binding antibodies and neutralisation

Our study population who received at least one dose of vaccine comprised 140 participants with a median age of 40 (interquartile range: 33, 48), 73 (52%) of whom were males. In order to analyse the proportion of participants in this urban population previously exposed to SARS-COV-2, we tested all baseline samples (n=140) for anti-N IgG using a flow cytometry based assay^18^ and found 62/140 participants were positive prior to administration of first vaccine dose, demonstrating 44% SARS-CoV-2 anti-N IgG seroprevalence at baseline prior to vaccination. Of the 62 subjects at baseline who were anti-N IgG positive, 12 became anti-N IgG negative at 1 month post second-dose and no further subjects lost anti-N IgG positivity between 1- and 3-months post second-dose. Of note, one subject who was anti-N IgG positive at baseline, became anti-N IgG negative at 1 month and then became anti-N IgG positive 3 month post second-dose with a 7-fold increase in anti-N IgG titres between 1 month and 3 months post second-dose - strongly suggestive of re-infection.

Of the 140 participants recruited (Figure 1), 49 had plasma samples available at baseline prior to vaccination and at two follow-up timepoints post vaccination for neutralization assays (Table 1). Median age was 39 (31, 46) and 47% were male. Half of the participants, 25/49 (51%) were IgG anti-N positive at baseline, and the geometric mean titre (GMT) of neutralizing antibodies associated with 50% neutralization (ID_50_) against WT PV across the entire study population was 145 ± 4.5(GMT±s.d) (Table 1; Figure 2a) with significantly lower titres observed against the Delta and Omicron variants with GMT titres 75 ± 3.6(GMT±s.d) and 55 ± 3.0(GMT±s.d) (p=0.0001 and p<0.0001) respectively. Baseline GMT of ID50s in the study population when stratified by anti-N status was 431 vs 47 in IgG anti-N positive and negative participants respectively, suggestive of the presence of neutralizing antibodies against SARS-CoV-2 in subjects negative for SARS-CoV2 anti-N Ab prior to vaccination. Of the 24 individuals anti-N Ab negative at baseline, 12/24 had ID_50_ above the cut-off of 20. In these individuals, binding antibodies to S were also detectable, and neutralisation correlated with IgG anti-S and IgG anti-RBD levels (r=0.71 and r=0.73) respectively indicating prior infection in at least half of those who were N Ab negative at baseline (Supplementary Figure 1).

**Table 1:** **Baseline characteristics of study participants** in longitudinal vaccine response study stratified by IgG anti-N status. ^a^ Serum Geometric mean titre (GMT) against Wuhan-614G wild type virus

|  |  | Follow-up |  |
| --- | --- | --- | --- |
|  | All | N-Ab +ve at baseline | N- Ab -ve at baseline |
| Characteristic |  |  |  |
| Total number, n (%) | 49 (100) | 25 (51) | 24 (49) |
| Age, median years (IQR) | 39 (31, 46) | 37 (31,44) | 40 (30, 49) |
| Male gender, n (%) | 23 (47) | 11 (44) | 12 (50) |
| Serum ID50 (GMT, 95 % CI) <sup>a</sup> |  |  |  |
| Baseline | 145 (95, 224) | 431 (297, 627) | 47 (29, 75) |
| 1 month post 2<sup>nd</sup> dose | 2579 (1689, 3938) | 2738 (4674, 7981) | 786 (1423, 2577) |
| 3 months post 2<sup>nd</sup> dose | 1112 (1695, 2584) | 1365 (2217, 3600) | 612 (1267, 2620) |

**Figure 2:**
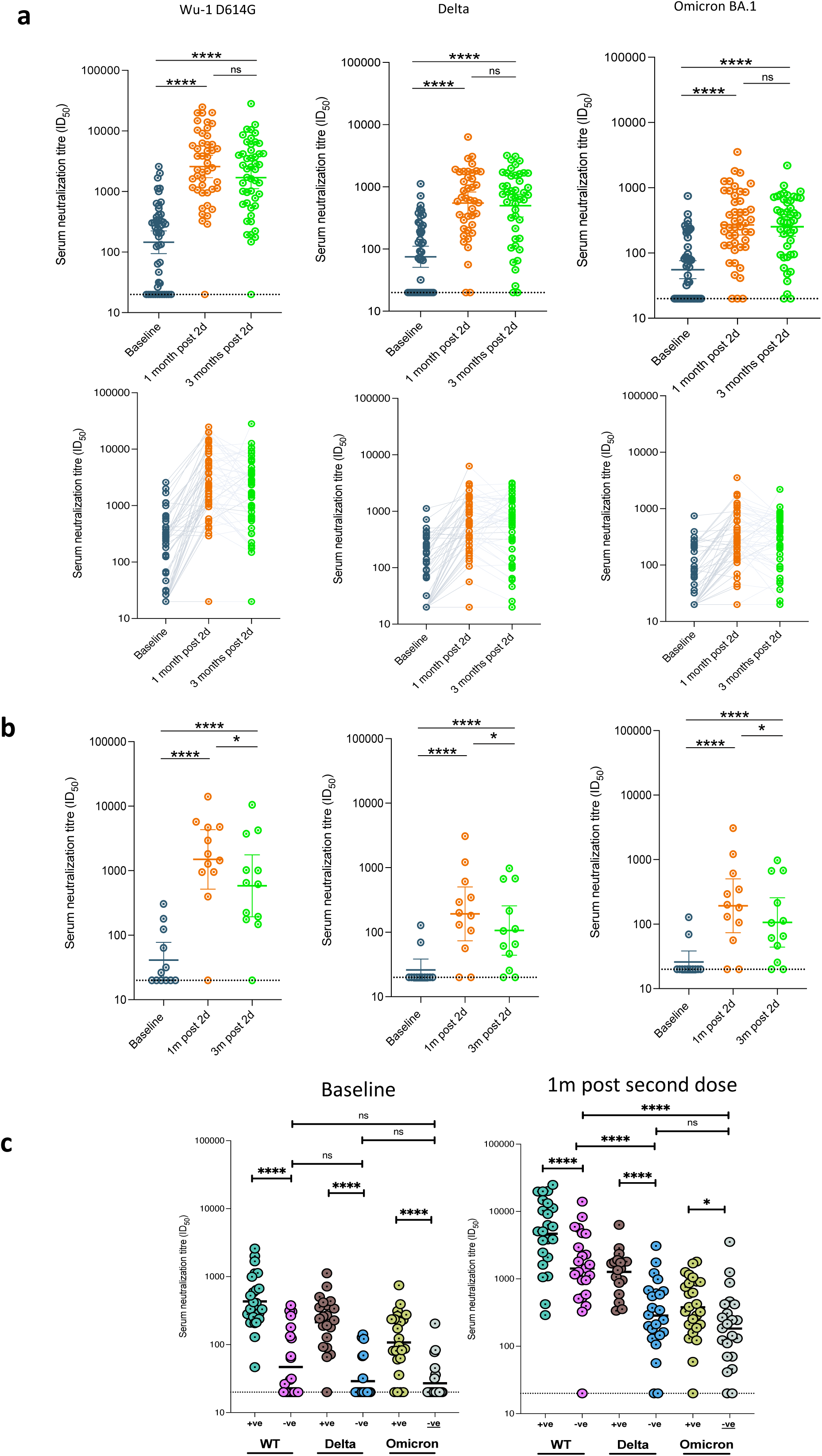
Longitudinal SARS-CoV-2 neutralization by sera from AZD1222 vaccinated individuals. **a)** Plasma neutralization of PV after two doses of the AZD1222 from participants at three consecutive time points: baseline (prior to first dose vaccination), 1mth after 2^nd^ dose vaccination and 3mth after vaccination (n=49). Data are representative of two independent experiments comprising of two technical replicates. **b)** Plasma neutralization of pseudotyped virus after two doses of the AZD1222 vaccine against VOC from (n=15) participants at baseline (prior to 1st dose vaccination), 1m (1 month) after 2nd dose vaccination and 3m (3 months) after vaccination and were anti-N IgG negative throughout study period. c) neutralisation titres before and after vaccine (n=49) stratified by N at baseline. Data shown as geometric mean titre (GMT) with 95% CI. Data are representative of two independent experiments comprising of two technical replicates. *P<0.05; **P<0.01; ***P<0.001; ****P<0.0001; ns=not significant.

### Neutralising and binding antibody responses following vaccination

Overall, neutralizing Ab titres to WT 1 month after second dose were 2579 ± 4.2(GMT±.s.d), and at 3 months post second-dose were 1695± 4.3(GMT±.s.d); p=0.07). As expected, lower levels of neutralization were observed against the Delta [549 ± 3.7(GMT±.s.d); p<0.0001)] and Omicron variants [269 ± 3.4(GMT±.s.d); p<0.0001] at 1 month, representing a fold reduction of 4.7 and 9.6 respectively (Figure 2). The GMT for Delta and Omicron was only around 100, nearly a log lower in comparison to WT (Figure 2). Positive anti-N IgG Ab status at baseline was associated with significantly higher titres of neutralizing antibodies following vaccination across all tested VOC (Figure 2). Those with anti-N Abs present at baseline did not experience waning of responses between months 1 and 3 post second dose (Supplementary Figure 2).

Overall, there was no decline in neutralising antibody titres at 3 months for WT, Delta, or and Omicron compared to 1 month post vaccination (Figure 2a). By contrast, when data were stratified by anti-N IgG status at any timepoint, there was a significant decline in neutralization between 1 month and 3 months post second-dose across all variants tested for participants who were N antibody negative throughout (Figure 2b; p=0.04). The GMT in these individuals for Delta and Omicron was approximately 100, nearly a log lower in comparison to WT (Figure 2B). Participants with anti-N Abs present at baseline did not experience waning of responses between months 1 and 3 post second dose (Supplementary Figure 2), despite frequent loss of N antibody over time (Supplementary Figure 3). When we examined binding antibodies over time in the group as a whole, we saw very small decreases for Wu-1 and Omicron Spike IgG but not for Wu-1 RBD (Supplementary Figure 4a). When we analysed the data for those N antibody negative, waning of binding antibodies was more evident (Supplementary Figure 4b, Supplementary Figure 5).

### Vaccine breakthrough infection

To evaluate the proportion of participants with vaccine breakthrough infection after two doses of AZD1222 vaccine, we tested anti-N IgG in subjects who were anti-N IgG negative at baseline (n=78) and became positive between 1- and 3-months post second dose and found 7/49 (14%) with *de-novo* infection, with one additional participant demonstrating both reinfection and breakthrough infection to yield a total breakthrough rate of 8/49 (16%, Figure 3 and Supplementary Figure 5). These individuals also experienced increase in antibodies to S and RBD that mirrored N antibody dynamics (Figure 3). We were also able to measure binding antibodies to Omicron that were around a log lower in titre as compared to Wu-1 binding antibodies as expected (Figure 3).

**Figure 3:**
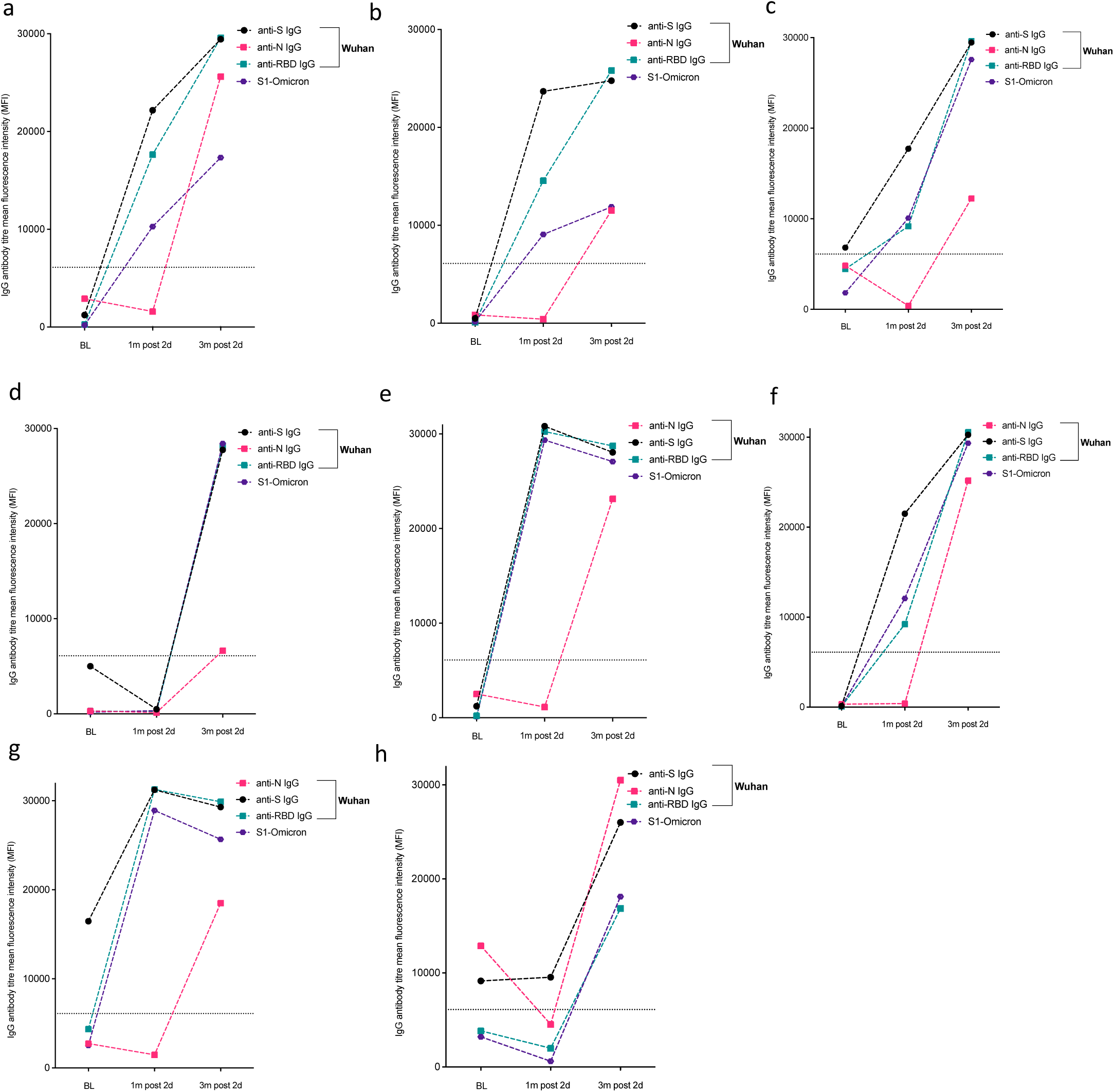
Kinetics of anti-SARS-COV-2 IgG antibodies in eight participants with evidence of breakthrough infection. (a-g), including one (h) with evidence of breakthrough infection and re-infection with SARS-COV-2 reinfection following evidence of positive IgG anti-N at baseline; negative IgG anti-N at 1 month after second dose and positive IgG anti-N 3 months post-second dose. Binding antibodies to Wu-1 and Omicron BA.1 are shown.

To investigate whether suboptimal immune response was related to subsequent breakthrough, we compared the neutralizing antibody titres 1 month post second dose between those with (n=8) or without breakthrough infection (n=15). We found no significant difference in neutralization between the groups (p=0.36, Figure 4a left panel). However, and as expected, neutralizing titres were higher at the last time point in individuals who had experienced vaccine breakthrough infection (with no evidence of infection prior to vaccine), indicating a boosting effect of infection in addition to vaccine (Figure 4a right panel). We noted that the increase in titres against Delta PV observed in breakthrough was significantly greater than the increase for WT and Omicron PVs, coincident with the Delta wave of infection in mid 2021.

**Figure 4:**
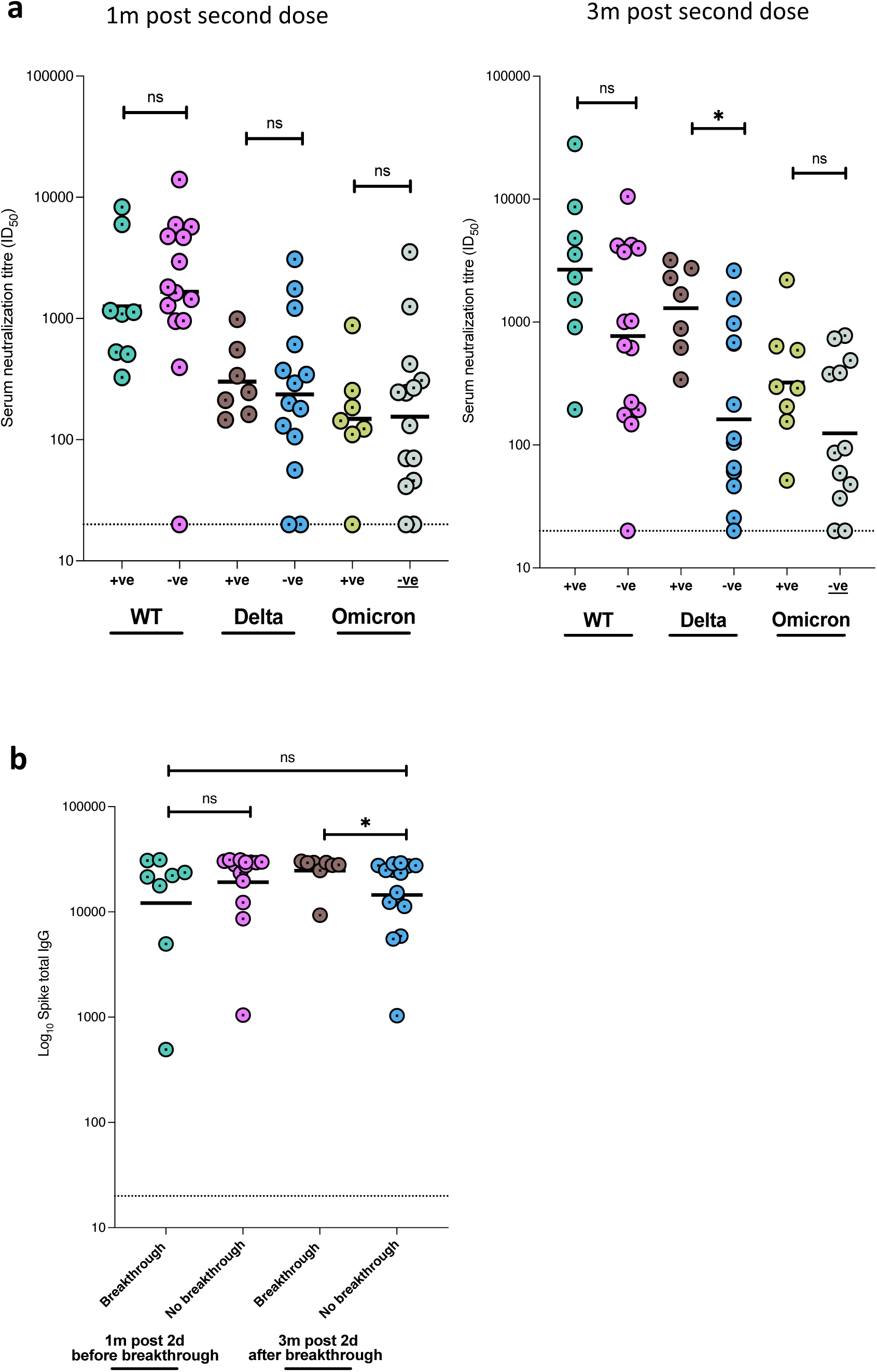
Neutralising and binding SARS-CoV-2 antibody responses one and three months after vaccination in context of breakthrough. **a**) Serum neutralization against pseudotyped virus (PV) from individuals with vaccine breakthrough occurring between months 1 and 3 post second dose and with no evidence of previous infection prior to vaccine (n=8, +ve), and those without breakthrough or past infection prior to vaccine (n=15, -ve). **b)** Total anti-spike binding IgG levels in individuals with breakthrough infection between 1 and 3 months post vaccination (n=8) and with no evidence of ‘natural’ infection (n=15).

## Discussion

A pivotal clinical trial for AZD1222 in South Africa (n=1010 vaccinees) during the Beta wave in mid to late 2020 showed poor protection against infection with Beta, correlating with immune escape *in vitro* using 19 samples from the vaccine arm^20^. No cases of severe disease or deaths were reported in the placebo or vaccine arms. Since then, AZD1222 has been deployed in more countries than any other vaccine (https://ourworldindata.org/covid-vaccinations), yet there are no real world data on two dose AZD1222 neutralising antibody responses from Africa. In addition, neither the impact of prior infection nor the impact of infection following vaccination with two dose AZD1222 on neutralising antibody responses have been reported in this setting. One study from Malawi in the pre Omicron era showed that a single dose of AZD1222 boosted neutralising and binding S antibody responses at 35 days post vaccine in 12 individuals with prior infection. There were no data on waning following vaccination or data on individuals not previously infected^21^.

Here, we first observed high prevalence (44%) of prior SARS-CoV-2 infection in Nigerian HCWs presenting for vaccination in early-2021, as determined by binding anti-N antibodies. There are no population-level SARS-CoV-2 seroprevalence data from Nigeria, but a recent modelling derived estimate of 72% in November 2021 was reported in a global analysis^22^. Anti-N Ab titres in some HCWs had declined to below cut-off in our Ab binding assay and detectable neutralisation and anti-S Ab in baseline pre-vaccine samples provided evidence of even higher prevalence of prior infection in this cohort prior to vaccination (>50%). It is clear that anti-N IgG measurements alone will miss some past infections requiring our combination approach to accurately measure prevalence. Notably, there was no vaccine available in country during the baseline screening period, and travel abroad was highly restricted. This rules out the possibility that those N negative and S positive had been vaccinated. Although waning of N antibody with rising titre upon re-infection has been reported before^23,24^, to our knowledge the ‘occult’ past infection revealed by neutralisation and presence of S binding antibodies has not been described in the African setting.

Our second major finding is that two AZD1222 vaccine doses led to a significant increase in neutralisation of WT Wu-1 D614G, Delta and Omicron PV, with GMT in the region of 1000 and higher titres in those who had evidence for prior infection at baseline. Data from high income settings have also shown a similar phenomenon^25,26^. A report from South Africa with single dose Ad26.CoV2.S also demonstrated vaccine boosting of infection-acquired immune responses^27^, however, unlike our study, these cited studies were conducted before emergence of the highly immune evasive Omicron variant^3^.

Thirdly, 16% of participants experienced infection between month 1 and month 3 post vaccination. Neutralising and binding Ab titres 1 month post vaccine, prior to breakthrough, did not appear to be associated with breakthrough infection, although this should be interpreted with caution given the small numbers and the fact that the community/occupational exposure to infection may have been heterogeneous. Those individuals with breakthrough had significantly higher plasma neutralisation for Delta PV but the differences for WT and Omicron PVs did not reach statistical significance. This is in keeping with the fact that Delta was circulating in summer of 2021^10,23^, and the breakthroughs were likely with Delta, explaining the specific increase in neutralisation against this variant. Binding Abs to WT and Omicron S protein increased significantly in those with breakthrough infection, though neutralising GMT for Omicron, whilst higher than in the non-infected, was nonetheless low in this group.

The limitations of the study include a modest sample size and follow up period, though we had nearly 50 participants with sequential follow up data and samples and 140 baseline samples for binding Ab studies. The underlying community population of Lagos was not sampled in a systematic way given vaccine delivery was first undertaken in individuals in the health sector; therefore the findings may not be fully generalisable to the whole country or region. In addition we did not measure non-neutralising antibody activities in vaccinees. Furthermore we were not able to identify the variants causing breakthrough infections, although neutralisation profiling was consistent with breakthrougxh infections largely driven by Delta.

We conclude that AZD1222 is immunogenic in this real world west African cohort with significantly higher than previously expected background seroprevalence and incidence of breakthrough infection over a short time period. Prior infection and breakthrough infection induced higher anti-SARS-CoV-2 Ab responses at 3 months post vaccine against all widely circulating VOC. However, plasma neutralisation against Omicron BA.1 was low at three months regardless of prior exposure. Two dose AZD1222 has been showed to generate lower neutralisation titres and efficacy across different settings, in comparison to mRNA vaccines^2,3,28^. mRNA vaccine ‘booster’ third doses induce broader, potent responses against Omicron BA.1^3,29^ and reduce mortality in the elderly^30^. Therefore, booster dosing after AZD1222 with mRNA vaccine should be considered in the African setting, even after natural infection and hybrid immunity, as future variants may be more pathogenic compared to BA.1^3,31,32^ whilst maintaining immune evasion on a background of waning immunity.

## Data Availability

All data produced in the present study are available upon reasonable request to the authors

## Acknowledgements

We would like to thank staff and study participants at the Nigeria Institute of Medical Research, Federal Medical Centre. A.A. is supported by Africa Research Excellence Fund Research Development Fellowship (AREF-318-ABDUL-F-C0882). A.A and S.A.K are supported by Bill and Melinda Gates Foundation via the Phylogenetics and Networks for Generalised Epidemics in Africa (PANGEA) (grant number OPP1175094). R.K.G is supported by a Wellcome Trust Senior Fellowship in Clinical Science (WT108082AIA).

## Competing interests

R.K.G has received honoraria for educational activities from Janssen, Moderna, and GSK.

## Authors contribution

Study conception and design: AA, DO, GO, OE, RA, RA, DF, BLS and RKG; Data collection: DO, AS, QO, RA, GL, OO; Performed experiments: AA, SK, SE, LCG, RD; Data analysis: AA, SK, DF J.A, CO, FI, SE, LCG, RD, SHA and RKG; Data interpretation: AA, DO, RA, JA, AS, FI, RD and RKG; Manuscript preparation; AA and RKG wrote the first draft of the manuscript with the critical input of all co-authors. All authors reviewed the results and approved the final version of the manuscript.

---

**Supplementary Figure 1:**
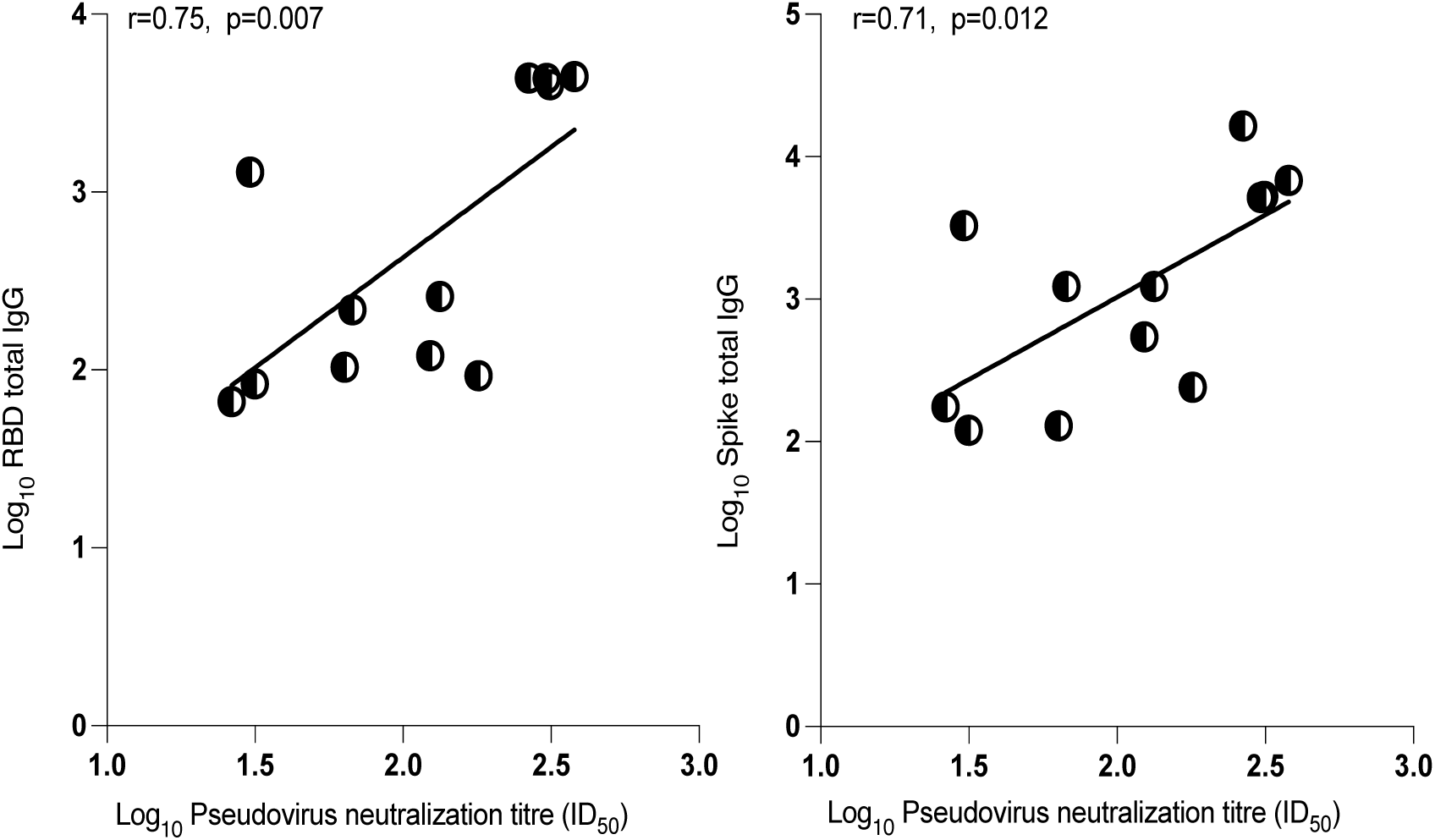
Spearman correlation between binding and neutralizing antibodies prior to vaccination. Correlation between anti-spike full length and receptor binding domain (RBD) IgG binding antibody responses and neutralization by sera against SARS-CoV-2 in a spike lentiviral pseudotyping assay expressing wild-type spike (D614G) **ns**=not significant. These data include only 11 individuals with ID50>20 at baseline and who were Anti-N antibody negative. ID50 is expressed as log_10_

**Supplementary Figure 2:**
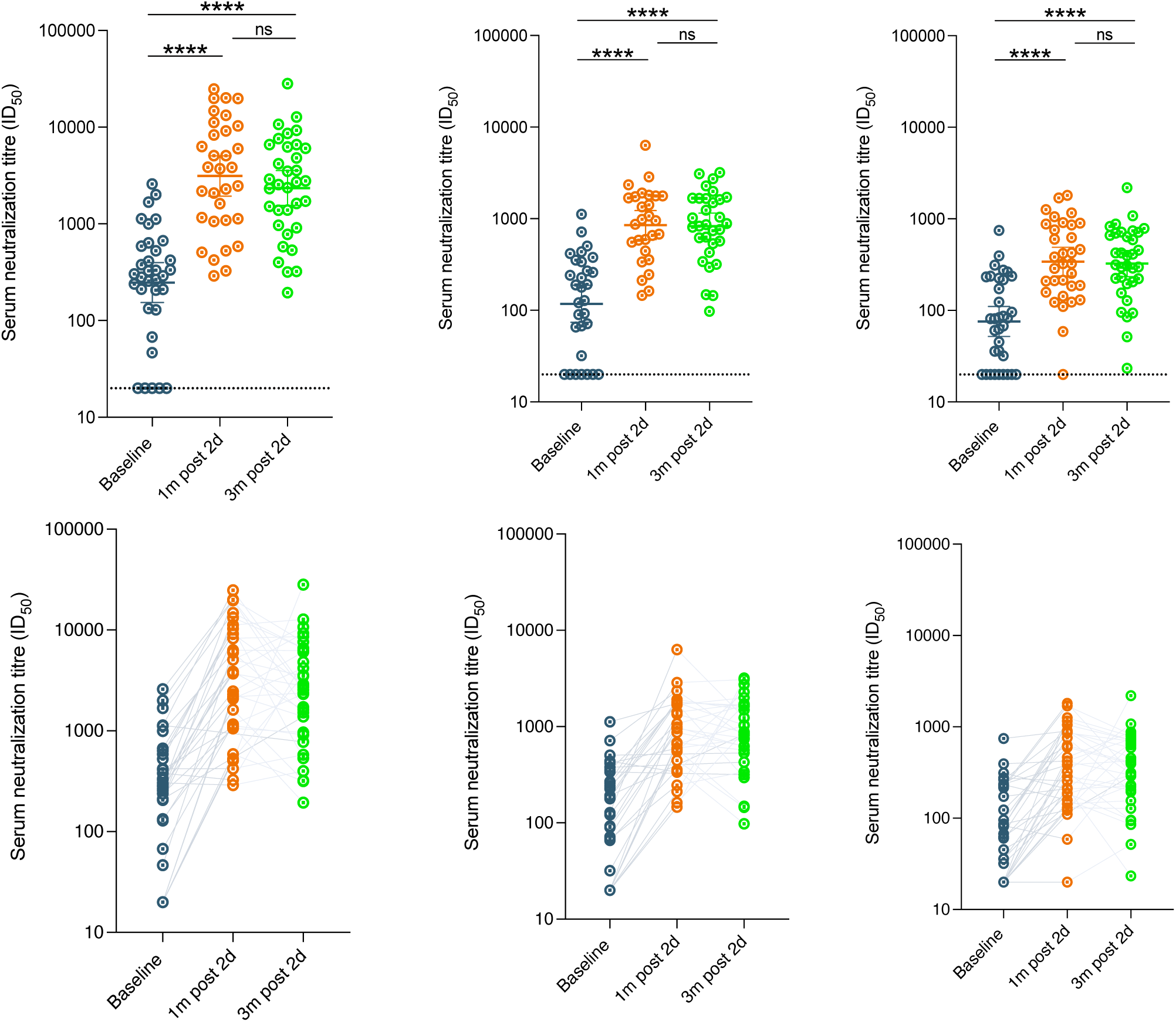
SARS-CoV-2 neutralization by sera from AZD1222 vaccinated individuals who were positive for N antibodies during follow up. Serum neutralization of pseudovirus after two doses of vaccine against pseudotyped virus (PV) expressing wild-type spike protein (D614G), delta and omicron variants of concerns from (n=34) participants at baseline (prior to first dose vaccination), 1m (1 month) after second dose vaccination and 3m (3 months) after vaccination and had ≥1 timepoint where anti-N IgG was positive on study. Data are representative of two independent experiments comprising of two technical replicates. Data shown as geometric mean titre (GMT) with 95% confidence interval. *P<0.05; ****P<0.0001; ns=not significant.

**Supplementary Figure 3:**
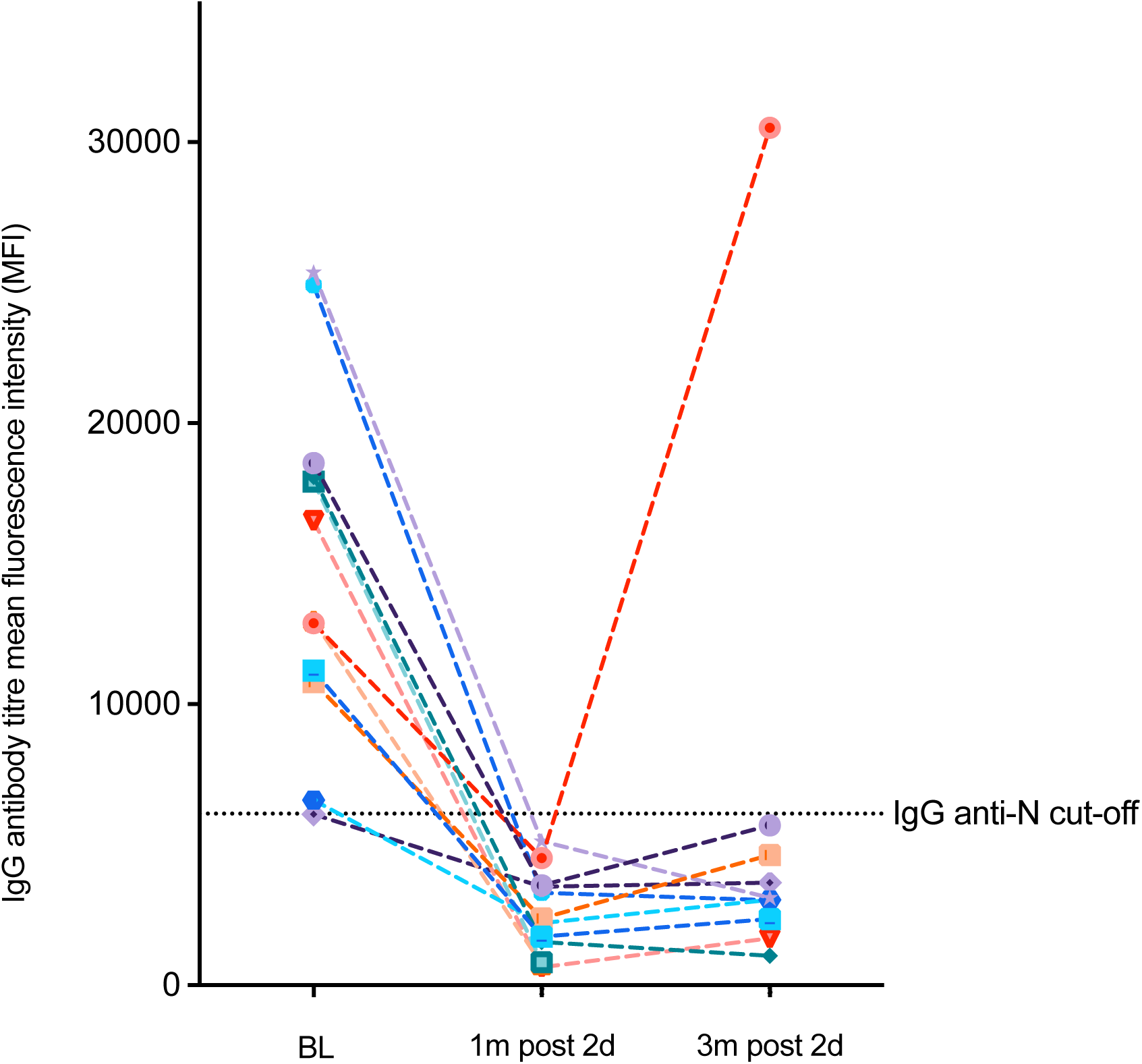
Kinetics of SARS-COV-2 total IgG anti-N antibodies in individuals positive for N at baseline. Note in orange one participant with SARS-COV-2 reinfection following evidence of positive IgG anti-N at baseline; negative IgG anti-N at 1 month after second dose and positive IgG anti-N 3 months post-second dose.

**Supplementary Figure 4:**
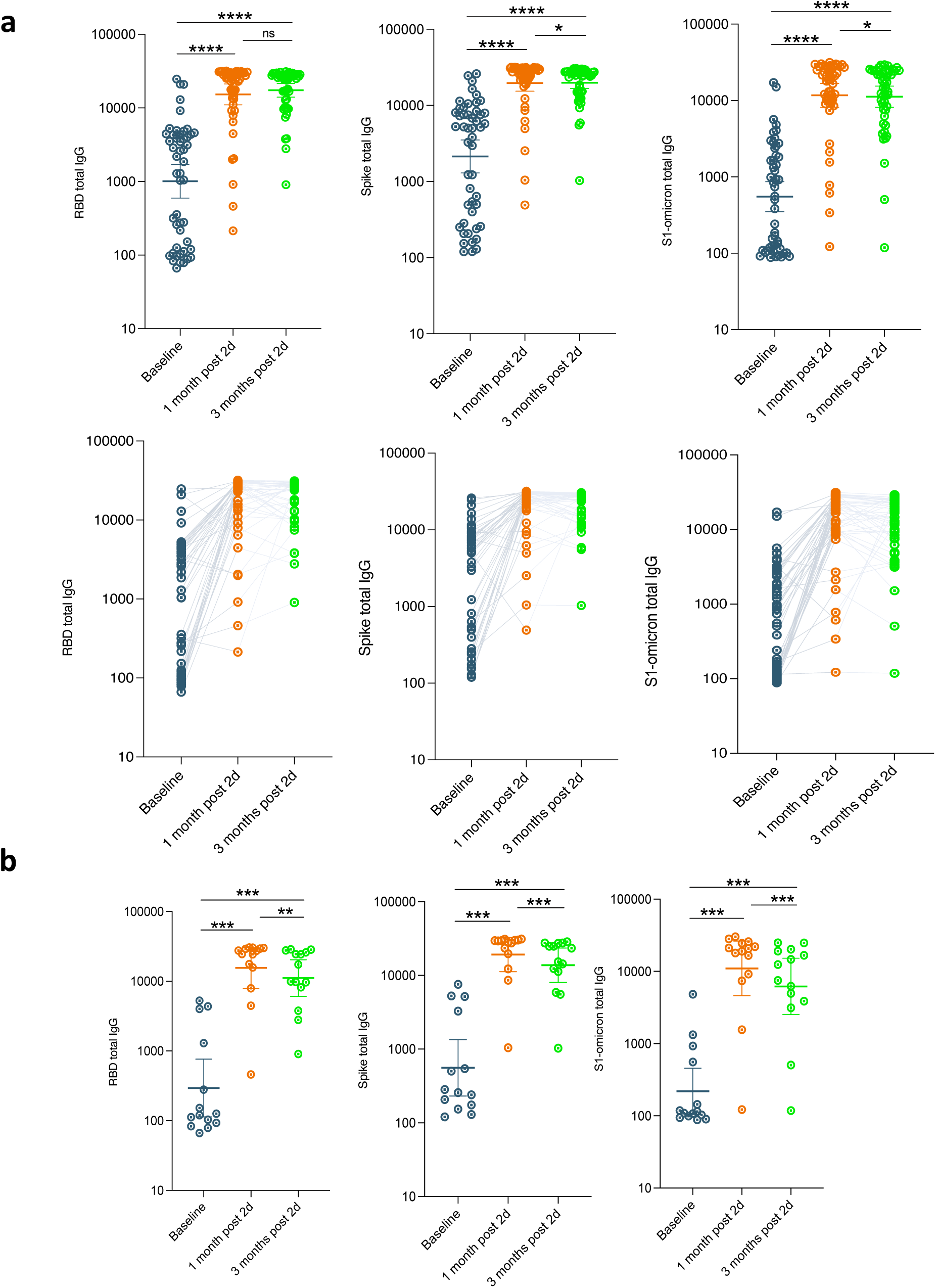
SARS-CoV-2 plasma binding antibodies against Wu-1 RBD and Spike S1 and Omicron BA.1 S1 from AZD1222 vaccinated individuals with 3 months follow up. **a)** Participants at baseline (prior to first dose vaccination), 1m (1 month) after second dose vaccination and 3m (3 months) after vaccination and had ≥1 timepoint where anti-N IgG was positive on study (N=39). **b)** Plasma binding antibodies (n=15) participants at baseline (prior to 1st dose vaccination), 1m (1 month) after 2nd dose vaccination and 3m (3 months) after vaccination in participants who were anti-N IgG negative throughout study period. Data are representative of two independent experiments comprising of two technical replicates. Data shown as mean fluorescence intensity (MFI) with 95% confidence interval. *p<0.05; **p<0.01, ***P<0.001, ****p<0.0001; ns=not significant.

**Supplementary Figure 5:**
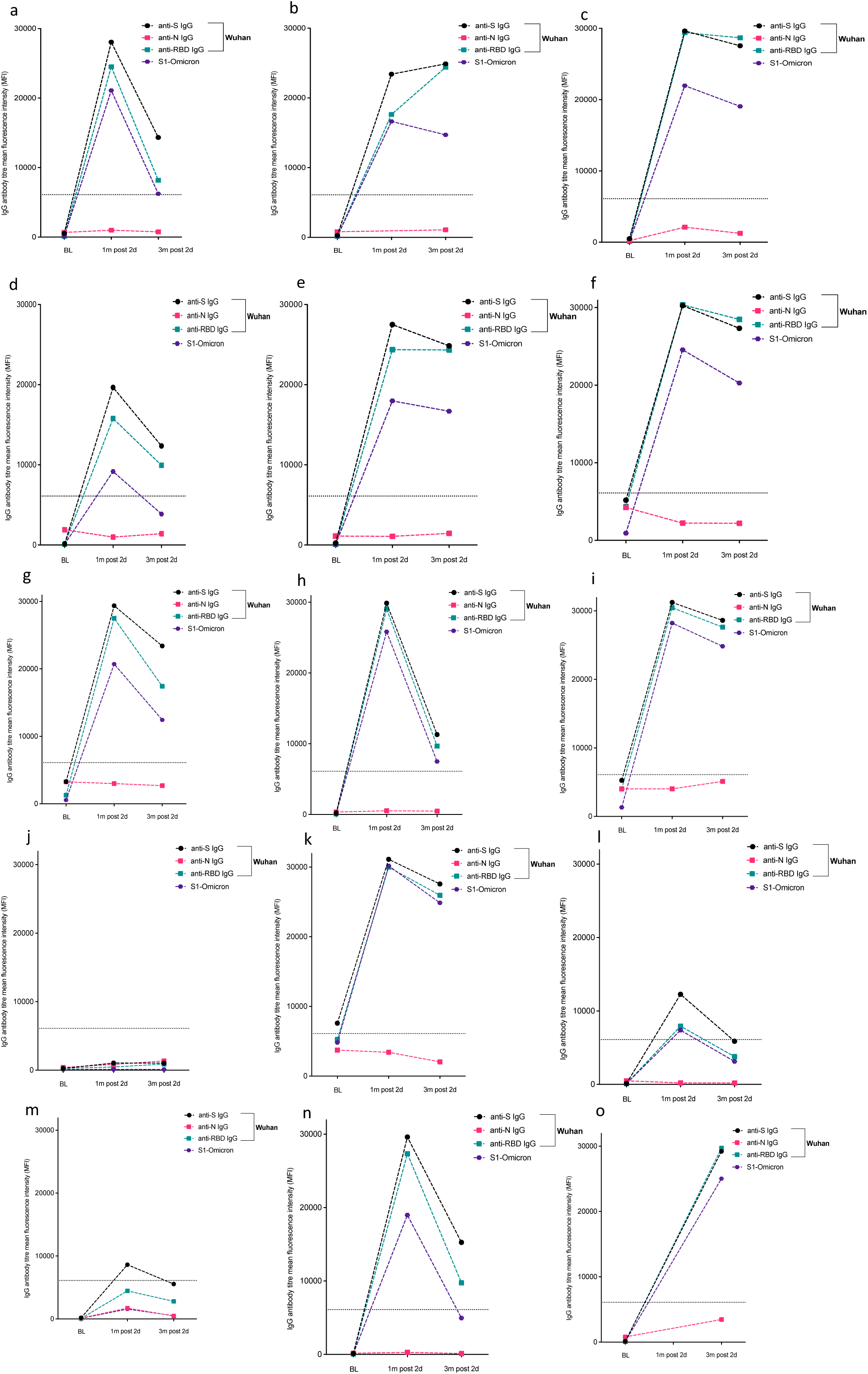
Kinetics of anti-SARS-COV-2 IgG binding antibodies to Wuhan-1 and Omicron in eight participants with no evidence of infection throughout study duration. (a-o), at baseline, 1 month after second dose and 3 months post-second dose. Binding antibodies to Wu-1 and Omicron BA.1 are shown. One subject (o) did not have sufficient sample volume available for testing at 1 month after second dose.

## Notes

### Competing Interest Statement

RG has received honoraria for educational activities from Janssen, Moderna, and GSK.

### Funding Statement

A.A. is supported by Africa Research Excellence Fund Research Development Fellowship AREF-318-ABDUL-F-C0882. R.K.G is supported by a Wellcome Trust Senior Fellowship in Clinical Science WT108082AIA.

### Author Declarations

This study was approved by the Institutional Review Board of National Institute of Medical Research, Nigeria IRB-21-040

### Summary of Updates

abstract altered to show dosing interval between doses of vaccine

